# A novel *SMARCC1*-mutant BAFopathy implicates epigenetic dysregulation of neural progenitors in hydrocephalus

**DOI:** 10.1101/2023.03.19.23287455

**Authors:** Amrita K. Singh, Stephen Viviano, Garrett Allington, Stephen McGee, Emre Kiziltug, Kedous Y. Mekbib, John P. Shohfi, Phan Q. Duy, Tyrone DeSpenza, Charuta G Furey, Benjamin C. Reeves, Hannah Smith, Shaojie Ma, André M. M. Sousa, Adriana Cherskov, August Allocco, Carol Nelson-Williams, Shozeb Haider, Syed R. A. Rizvi, Seth L. Alper, Nenad Sestan, Hermela Shimelis, Lauren K. Walsh, Richard P. Lifton, Andres Moreno-De-Luca, Sheng Chih Jin, Paul Kruszka, Engin Deniz, Kristopher T. Kahle

**Author notes:** <u>Contact:</u> Kristopher T. Kahle, M.D., Ph.D., Massachusetts General Hospital and Harvard Medical School, Department of Neurosurgery, 55 Fruit St., Wang Ambulatory Care Center, Suite 333, Boston, MA 02114, USA.

## Abstract

**Importance:** Hydrocephalus, characterized by cerebral ventriculomegaly, is the most common disorder requiring brain surgery. A few familial forms of congenital hydrocephalus (CH) have been identified, but the cause of most sporadic cases of CH remains elusive. Recent studies have implicated *SMARCC1*, a component of the <u>B</u>RG1-<u>a</u>ssociated factor (BAF) chromatin remodeling complex, as a candidate CH gene. However, *SMARCC1* variants have not been systematically examined in a large patient cohort or conclusively linked with a human syndrome. Moreover, CH-associated *SMARCC1* variants have not been functionally validated or mechanistically studied *in vivo*.

**Objectives:** The aims of this study are to (i) assess the extent to which rare, damaging *de novo* mutations (DNMs) in *SMARCC1* are associated with cerebral ventriculomegaly; (ii) describe the clinical and radiographic phenotypes of *SMARCC1*-mutated patients; and (iii) assess the pathogenicity and mechanisms of CH-associated *SMARCC1* mutations *in vivo*.

**Design, setting, and participants:** A genetic association study was conducted using whole-exome sequencing from a cohort consisting of 2,697 ventriculomegalic trios, including patients with neurosurgically-treated CH, totaling 8,091 exomes collected over 5 years (2016-2021). Data were analyzed in 2023. A comparison control cohort consisted of 1,798 exomes from unaffected siblings of patients with autism spectrum disorder and their unaffected parents sourced from the Simons simplex consortium.

**Main outcomes and measures:** Gene variants were identified and filtered using stringent, validated criteria. Enrichment tests assessed gene-level variant burden. *In silico* biophysical modeling estimated the likelihood and extent of the variant impact on protein structure. The effect of a CH-associated *SMARCC1* mutation on the human fetal brain transcriptome was assessed by analyzing RNA-sequencing data. *Smarcc1* knockdowns and a patient-specific *Smarcc1* variant were tested in *Xenopus* and studied using optical coherence tomography imaging, *in situ* hybridization, and immunofluorescence microscopy.

**Results:** *SMARCC1* surpassed genome-wide significance thresholds in DNM enrichment tests. Six rare protein-altering DNMs, including four loss-of-function mutations and one recurrent canonical splice site mutation (c.1571+1G>A) were detected in unrelated patients. DNMs localized to the highly conserved DNA-interacting SWIRM, Myb-DNA binding, Glu-rich, and Chromo domains of *SMARCC1*. Patients exhibited developmental delay (DD), aqueductal stenosis, and other structural brain and heart defects. G0 and G1 *Smarcc1 Xenopus* mutants exhibited aqueductal stenosis and cardiac defects and were rescued by human wild-type *SMARCC1* but not a patient-specific *SMARCC1* mutant. Hydrocephalic *SMARCC1*-mutant human fetal brain and *Smarcc1*-mutant *Xenopus* brain exhibited a similarly altered expression of key genes linked to midgestational neurogenesis, including the transcription factors *NEUROD2* and *MAB21L2*.

**Conclusions:** *SMARCC1* is a *bona fide* CH risk gene. DNMs in *SMARCC1* cause a novel human BAFopathy we term “<u>S</u>MARCC1-<u>a</u>ssociated <u>D</u>evelopmental <u>D</u>ysgenesis <u>S</u>yndrome (SaDDS)”, characterized by cerebral ventriculomegaly, aqueductal stenosis, DD, and a variety of structural brain or cardiac defects. These data underscore the importance of SMARCC1 and the BAF chromatin remodeling complex for human brain morphogenesis and provide evidence for a “neural stem cell” paradigm of human CH pathogenesis. These results highlight the utility of trio-based WES for identifying risk genes for congenital structural brain disorders and suggest WES may be a valuable adjunct in the clinical management of CH patients.

**KEY POINTS:** *Question:* What is the role of *SMARCC1*, a core component of the <u>B</u>RG1-<u>a</u>ssociated factor (BAF) chromatin remodeling complex, in brain morphogenesis and congenital hydrocephalus (CH)?

*Findings:* *SMARCC1* harbored an exome-wide significant burden of rare, protein-damaging *de novo* mutations (DNMs) (p = 5.83 × 10^−9^) in the largest ascertained cohort to date of patients with cerebral ventriculomegaly, including treated CH (2,697 parent-proband trios). *SMARCC1* contained four loss-of-function DNMs and two identical canonical splice site DNMs in a total of six unrelated patients. Patients exhibited developmental delay, aqueductal stenosis, and other structural brain and cardiac defects. *Xenopus Smarcc1* mutants recapitulated core human phenotypes and were rescued by the expression of human wild-type but not patient-mutant *SMARCC1*. Hydrocephalic *SMARCC1*-mutant human brain and *Smarcc1*-mutant *Xenopus* brain exhibited similar alterationsin the expression of key transcription factors that regulate neural progenitor cell proliferation.

*Meaning:* *SMARCC1* is essential for human brain morphogenesis and is a *bona fide* CH risk gene. *SMARCC1* mutations cause a novel human BAFopathy we term “<u>S</u>MARCC1-<u>a</u>ssociated <u>D</u>evelopmental <u>D</u>ysgenesis <u>S</u>yndrome (SaDDS)”. These data implicate epigenetic dysregulation of fetal neural progenitors in the pathogenesis of hydrocephalus, with diagnostic and prognostic implications for patients and caregivers.

## INTRODUCTION

Epigenetic mechanisms, including methylation, histone modifications, and ATP-dependent chromatin remodeling, regulate gene expression by altering chromatin structure.^1-4^ The SWI/SNF (SWItch/Sucrose Non-Fermentable) complex (also known as the BRG1-associated factor [BAF] complex) is one of four ATP-dependent chromatin remodeling complexes known in mammals.^5-7^ The BAF complex mediates nucleosome modification critical to modulating gene expression in multiple essential processes, including cell differentiation and proliferation, and DNA repair.^1-5^ The combinatorial assembly of numerous gene family paralogs yields many potential types of complex hetero-oligomeric complexes that provide tissue and temporal specificity^6-8^ for the control of gene transcription that is essential for the development of the brain,^9,10^ heart,^11-13^ and other organs, as well as the maintenance of embryonic stem cell pluripotency (Table 1).^14^

**TABLE 1.**
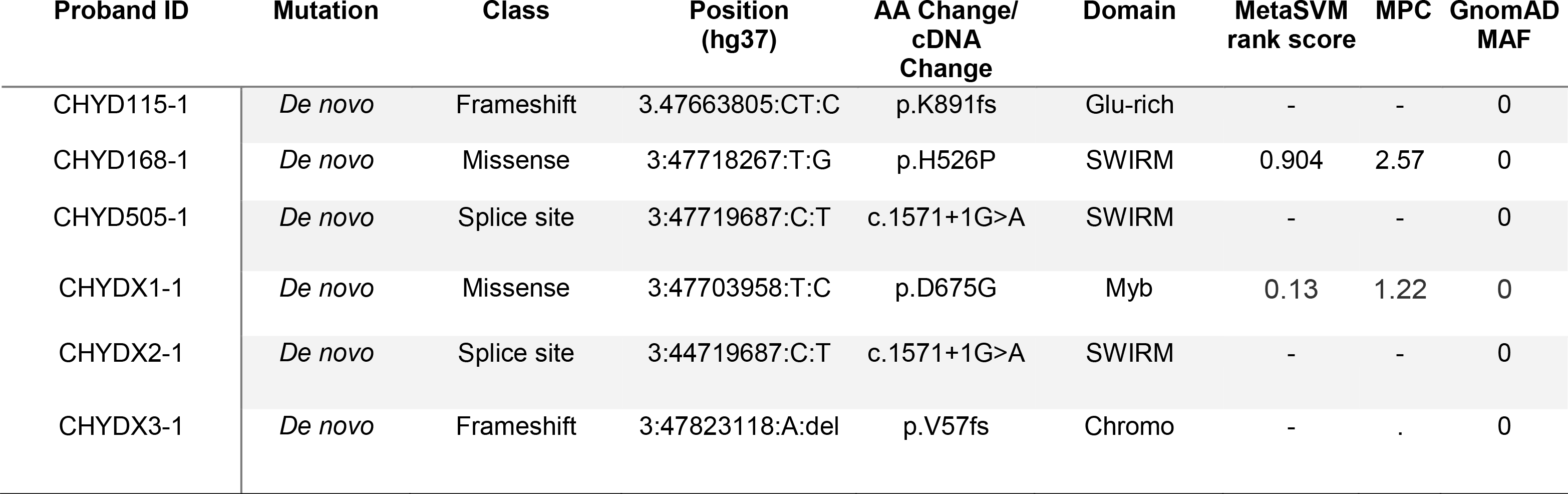
*De novo* mutations in *SMARCC1* probands.

BAFopathies constitute a heterogeneous group of disorders caused by variants in various subunits composing the BAF complex.^15^ The phenotypic spectrum of BAFopathies includes intellectual disability (ID) and developmental delay (DD), autism, schizophrenia, amyotrophic lateral sclerosis,^20-22^ and other human neurodevelopmental disorders and anatomical congenital defects.^16,17^ The most recognizable syndrome associated with BAF abnormalities is Coffin-Siris syndrome (CSS [MIM: 135900]). This is a genetically heterogeneous ID/DD syndrome characterized by speech delay, coarse facial appearance, feeding difficulties, hypoplastic-to-absent fifth fingernails, and fifth distal phalanges. ^18^ This syndrome is associated with variants in multiple BAF complex subunits, including the ATPase subunit *SMARCA4* (MIM: 603254), the common core subunit *SMARCB1* (MIM: 601607), and BAF accessory subunits such as *SMARCE1/BAF57* (MIM: 603111), *ARID1A* (MIM: 603024), *ARID1B* (MIM: 614556), *ARID2* (MIM: 609539), and *DPF2* (MIM: 601671).^18,19^ Other BAFopathies, such as Nicolaides-Baraitser syndrome (MIM: 601358), have significant phenotypic overlap with CSS and are caused by pathogenic variants in *SMARCA2* (MIM: 600014).^20-22^

*SMARCC1* (SWI/SNF-Related, Matrix-Associated, Actin-Dependent Regulator Of Chromatin Subfamily C Member 1) encodes an essential core subunit of the BAF complex highly homologous to *SMARCC2*.^8,23^ *Smarcc1* is highly expressed in the mouse embryonic neuroepithelium and ventricular zone.^9,10,14^Similar to other components of the neuroprogenitor-specific BAF complexes, Smarcc1regulates the proliferation, differentiation, and survival of mouse neural progenitors via transcriptional regulation of genes critical for telencephalon development.^24-27^ *Smarcc2; Smarcc1* double knockout mice exhibit proteasome-mediated degradation of the entire BAF complex, resulting in impairment of the global epigenetic and gene expression program of cortical development.^15,28^ *Smarcc1* knockout causes embryonic lethality in mice.^29,30^ ∼80% of mice homozygous for the *Smarcc1*^*msp/msp*^ missense allele exhibit exencephaly due to decreased proliferation and increased apoptosis of neural progenitors in the neural tube.^29,31^

Recently, whole-exome sequencing (WES) studies in patients with congenital hydrocephalus (CH) identified *SMARCC1* as a candidate gene, implicating impaired epigenetic regulation of neural progenitor cell (NPC) proliferation and differentiation in the development of ventriculomegaly.^32,33^ However, despite its significant biological role, *SMARCC1* variants have not been conclusively associated with a human syndrome, and CH-associated *SMARCC1* variants have been neither functionally assessed nor mechanistically studied *in vivo*. The objectives of this study were to: (i) assess the extent to which rare, damaging *de novo* mutations (DNMs) in *SMARCC1* are associated with CH risk; (ii) describe the phenotypes of *SMARCC1*-mutant patients; and (iii) functionally-validate and assess the cellular and molecular mechanisms of CH-associated *SMARCC1* mutation in a novel animal model of hydrocephalus.

Our findings show *SMARCC1* is a *bona fide* CH risk gene and suggest rare, damaging germline DNMs in *SMARCC1* cause a novel human BAFopathy we term “SMARCC1-associated Developmental Dysgenesis Syndrome (SaDDS)” characterized by DD, cerebral ventriculomegaly, and other structural brain or cardiac defects. Our data highlight the importance of the ATP-dependent BAF chromatin remodeling complex for human brain morphogenesis and CSF dynamics and further support a “neural stem cell paradigm” of human CH.^34-36^ These data highlight the power of trio-based WES for identifying pathogenic mutations in sporadic structural brain disorders and suggest its utility as a prognostic adjunct when evaluating the surgical candidacy and prognosis of CH patients.

## RESULTS

The sequenced cohort consisted of a total of 8,091 exomes that included an expanded cohort of 2,697 trios with cerebral ventriculomegaly, including 281 with neurosurgically-treated congenital hydrocephalus (CH) (see Methods). The control cohort consisted of 1,798 exomes from unaffected siblings of people diagnosed with autism spectrum disorder and their unaffected parents sourced from the Simons simplex consortium.^32,37^ Genomic DNAs were subjected to WES, and variant calling was performed with GATK HaplotypeCaller and Freebayes followed by ANNOVAR annotation and confirmation by the Integrative Genomics Viewer.^33,38,39^ Reported variants were confirmed by Sanger sequencing.

We compared observed and expected numbers of non-synonymous *de novo* variants (DNVs) in all genes in CH cases and controls (see Methods). *SMARCC1* surpassed genome-wide significance thresholds for DNV burden (multiple-testing correction threshold of 8.57 × 10^−7^ after correction for testing 19,347 RefSeq genes in triplicate using a one-tailed Poisson test (Figure 1a, Extended Data Table 1). *SMARCC1* harbored a total of six DNVs (p.His526Pro, p.Lys891fs, p.Asp675Gly, p.Val57fs, and the recurrent c.1571+1G>A splice site mutation) in unrelated patients (Table 1). The probands with p.His526Pro, p.Lys891fs, as well as one of the two probands with c.1571+G>A have been previously reported.^32,33^ p.His526Pro is predicted to abolish interaction with the backbone carbonyl oxygen of p.Leu505 at the end of an adjacent helix in the SWIRM domain mediating BAF complex subunit interactions.^40^ p.Asp675Gly alters a conserved residue in the Myb domain resulting in an unfavorable loss of an ion pair interaction with p.Arg602 (Extended Data Figure 1). All these DNVs are absent in gnomAD and Bravo databases. The probability of encountering six damaging DNVs in *SMARCC1* in a cohort of this size was calculated as 3.88 × 10^−15^, an exceedingly unlikely chance event. No significant enrichment of *SMARCC1* DNMs was observed in control trios.

**FIGURE 1.**
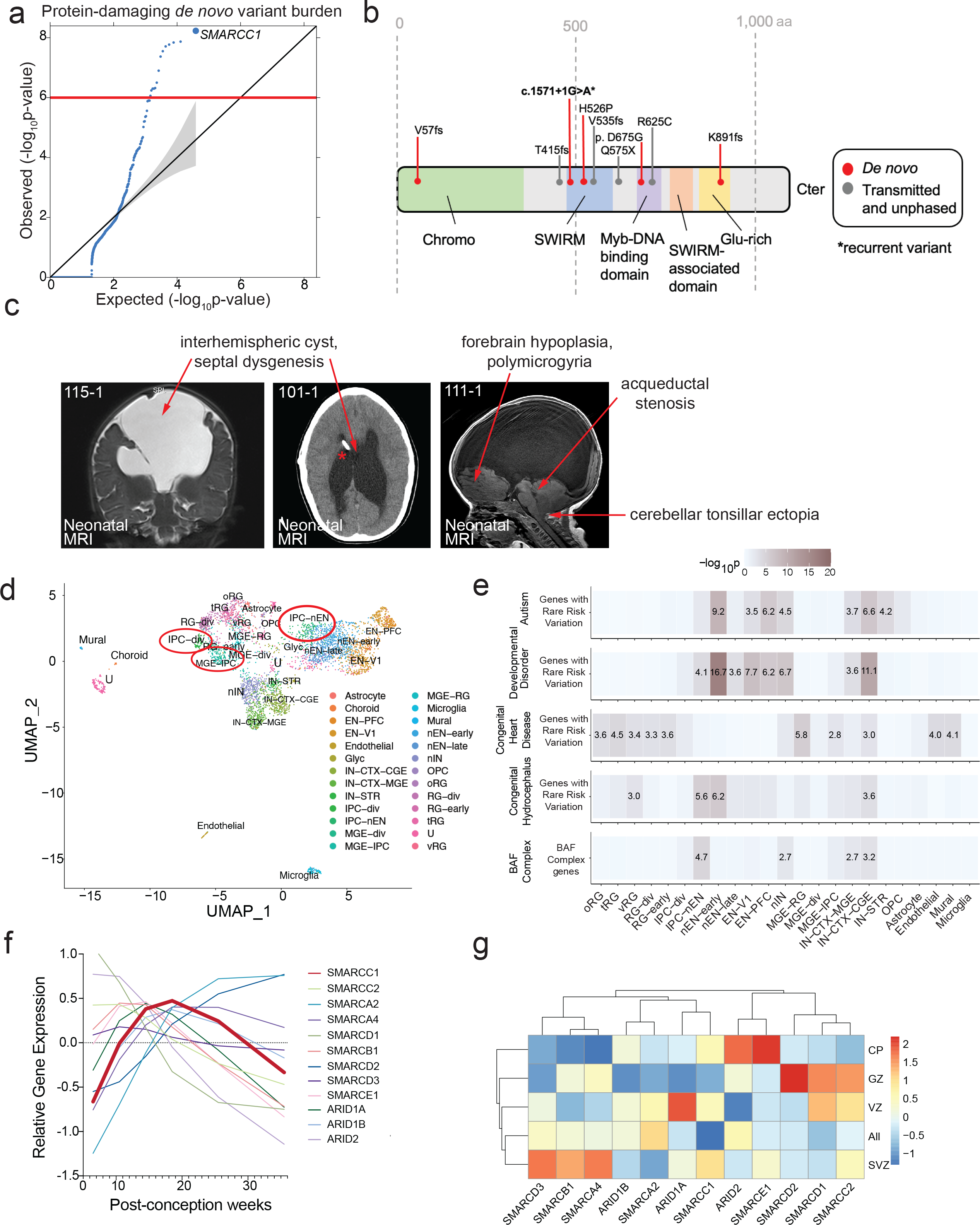
*SMARCC1* mutations are associated with congenital hydrocephalus (CH) and cause a novel human BAFopathy featuring cerebral ventriculomegaly. A. Quantile–quantile (Q-Q) plot of observed versus expected P-values for DNMs in each gene in 2,697 trio cases. P-values were calculated using a one-sided Poisson test (see Methods). For protein-damaging *de novo SMARCC1* variants (LoF, MetaSVM = D, and/or MPC > 2), p = 5.83 × 10^−9^. B. Schematic diagram showing variant locations in SMARCC1 protein domains. *De novo* variants are indicated with red markers, transmitted and unphased variants^32,33^ are indicated with gray markers. Asterisk indicates recurrent variant. C. Brain MRIs of CH patients with *SMARCC1* variants demonstrate consistent structural abnormalities. Prenatal imaging is shown for patients 115-1 (contrast MRI), 101-1, and 111-1. Red asterisks denote ventricular catheter of a ventriculo-peritoneal shunt used to treat obstructive hydrocephalus. D. Analyzed transcriptomic dataset^24^ showing MAP clustering of developmental human brain cells, colored by cell type. Early and late born excitatory neuron PFC (EN-PFC), early and late born excitatory neuron V1 (EN-V1), CGE/LGE-derived inhibitory neurons (IN-CTX-CGE), MGE-derived ctx inhibitory neuron (IN-CTX-MGE), striatal neurons (IN-STR), dividing intermediate progenitor cells RG-like (IPC-div), intermediate progenitor cells EN-like (IPC-nEN), dividing MGE progenitors (MGE-div), MGE progenitors (MGE-IPC), MGE radial glia (MGE-RG), mural/pericyte (Mural), newborn excitatory neuron - early born (nEN-early), newborn excitatory neuron - late born (nEN-late), MGE newborn neurons (nIN), oligodendrocyte progenitor cell (OPC), outer radial glia (oRG), dividing radial glia (RG-div), earlyvRG (RG-early), truncated Radial Glia (tRG), Unknown (U), Ventricular Radial Glia (vRG). Expression in neural progenitors is featured in red circles. E. Analyzed transcriptomic dataset^24^ showing enrichment analysis across cell type markers of the developmental human brain for genes with rare risk variation in autism, developmental disorders, congenital heart disease, and congenital hydrocephalus compared to BAF Complex Genes. Tiles labeled with −log_10_(*P* value) and an asterisk represent significant enrichment at the Bonferroni multiple-testing cutoff (α□=□0.05/23□=□2.17□×□10^−3^). F. Temporal gene expression profiles for *SMARCC1* and other BAF Complex genes between post-conception weeks (PCW) 5-36. G. Analyzed transcriptomic dataset^24^ showing heatmap of gene expression levels for for *SMARCC1* and other BAF complex genes across cortical lamina, PCW 5-40. CP, cortical plate; GZ, germinal zone; VZ, ventricular zone; SVZ, subventricular zone.

We examined the clinical phenotypes of probands harboring *SMARCC1* DNVs and other published rare, damaging transmitted or unphased CH-associated *SMARCC1* associated variants^33^ (Extended Data Table 2). The latter included two transmitted LoF mutations (p.Gln575X and p.Val535fs), one unphased rare LoF variant (p.Thr415fs), and one transmitted rare damaging missense (D-Mis) variant (p.Arg652Cys). Strikingly, 10/10 had perinatally diagnosed cerebral ventriculomegaly, and at least 7 required neurosurgical CSF diversion by endoscopic third ventriculostomy or ventriculoperitoneal shunting. 9/10 had aqueductal stenosis. 9/10 had partial or complete corpus callosum abnormalities, including septal agenesis. 9/10 exhibited moderate to profound DD. 9/10 had cardiac defects including atrial septal defect, ventricular septal defect, double outlet right ventricle, and cardiac hypoplasia. Other neurodevelopmental phenotypes, such as seizures, structural brain defects like cerebellar tonsillar ectopia, and craniofacial defects including cleft palate, microtia, and auditory canal atresia were variably present (Table 2, Figure 1c). These data suggest that *SMARCC1* mutation, in addition to conferring CH risk, leads to a novel human syndrome.

**TABLE 2.**
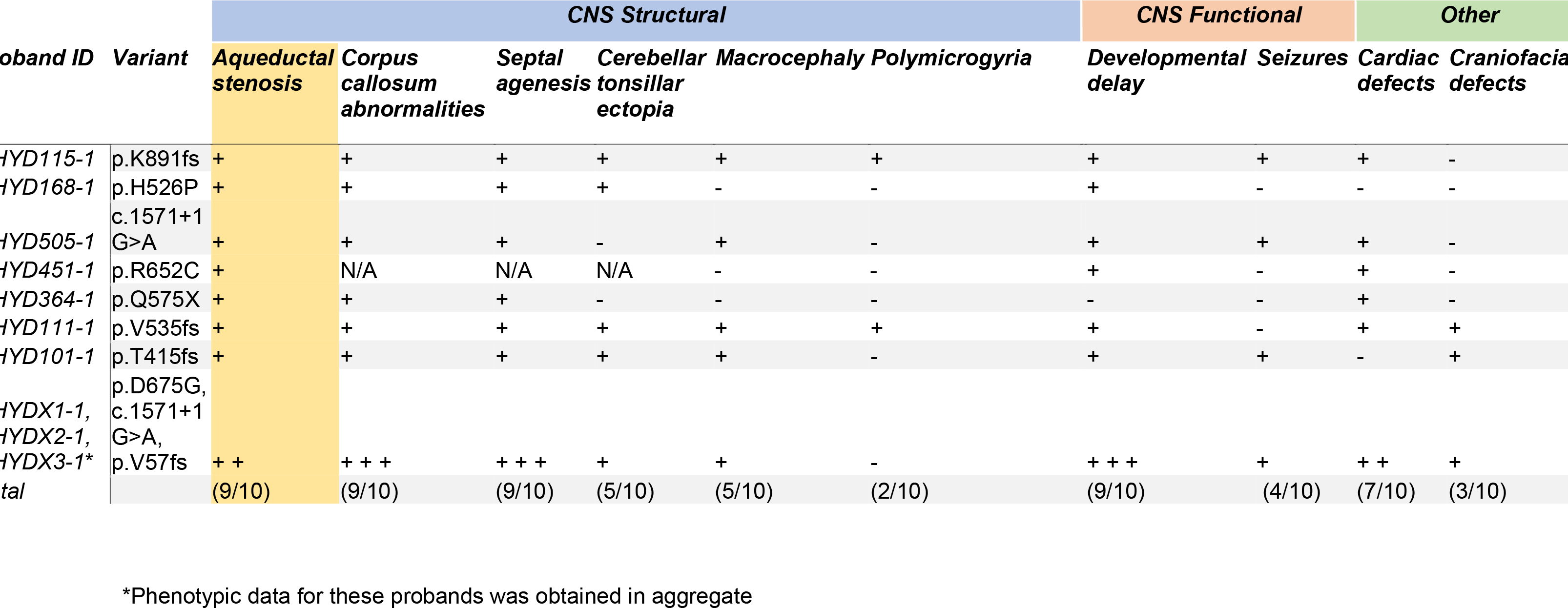
Phenotypic characteristics of *de novo* and transmitted SMARCC1 probands.

*Smarcc1* is expressed in mouse ventricular zone neuroepithelial and neural progenitor cells during midgestation,^10,32,41,42^ a key epoch during which neurogenesis contributes to the development of the diencephalon and telencephalon.^9,24,29,42-45^ We studied the expression of *SMARCC1* in the human brain during development using single-cell RNA-sequencing database of 4,261 cells from developmental human whole brain tissue during PCW 6-40.^24^ We found that *SMARCC1* and other BAF complex genes are expressed highly in intermediate progenitor cells (IPCs) between PCW 13-20. (Figure 1g, Extended Data Figure 2). In addition, *SMARCC1* is highly expressed in the lateral ganglionic eminence, a NPC niche within the ventral telencephalon that harbors NPCs destined for cortical and striatal interneurons and oligodendrocyte precursor cells (OPCs) (Extended Data Figure 2).^46^ When investigating ventricular lamina expression, we found that BAF complex members are also expressed throughout the ventricular laminae. Further, *SMARCC1* is most highly expressed in the ventricular zone (Figure 1g). These data show that SMARCC1 is highly expressed in human fetal periventricular NPCs.

To functionally validate *SMARCC1* as a novel disease gene, we generated *Smarcc1* mutant *Xenopus tropicalis* tadpoles since mice with *Smarcc1* deletion are embryonic lethal.^29-31^ Additionally, brain morphogenesis and CSF circulation can be studied in live *Xenopus* tadpoles using optical coherence tomography (OCT) (Extended Data Figure 3).^47,48^ We knocked down *Xenopus Smarcc1* using CRISPR/CAS9 targeting exon1 and exon10, as well as by using a morpholino oligo targeting the *Smarcc1* transcription start site (Figure 2a, 2b, Extended Data Figure 4). All three resulting *Smarcc1* mutant and morphant tadpoles exhibited highly penetrant aqueductal stenosis that was transmitted to G1 mutant progeny (Figure 2a, 2b). Despite this, OCT imaging demonstrated intact ependymal cilia-driven CSF circulation (Extended Data Figure 5). Overexpression of human wild-type *SMARCC1* but not human CH-variant *SMARCC1* p.Gln575X in *Smarcc1*-depleted *X. tropicalis* mutants (see Methods) rescued aqueductal stenosis (Figure 2c, Extended Data Figure 6). *Smarcc1*-depleted variants also exhibited decreased cardiac function resembling hypoplastic cardiomyopathy (e.g., see proband CHYD111-1), as quantified by end-diastolic diameter, end-systolic diameter, and shortening fraction (Extended Data Figure 7). These results show CH-associated *SMARCC1* mutation or *Smarcc1* depletion in *Xenopus* phenocopies the core brain and cardiac pathology of humans with *SMARCC1* mutations.

**FIGURE 2.**
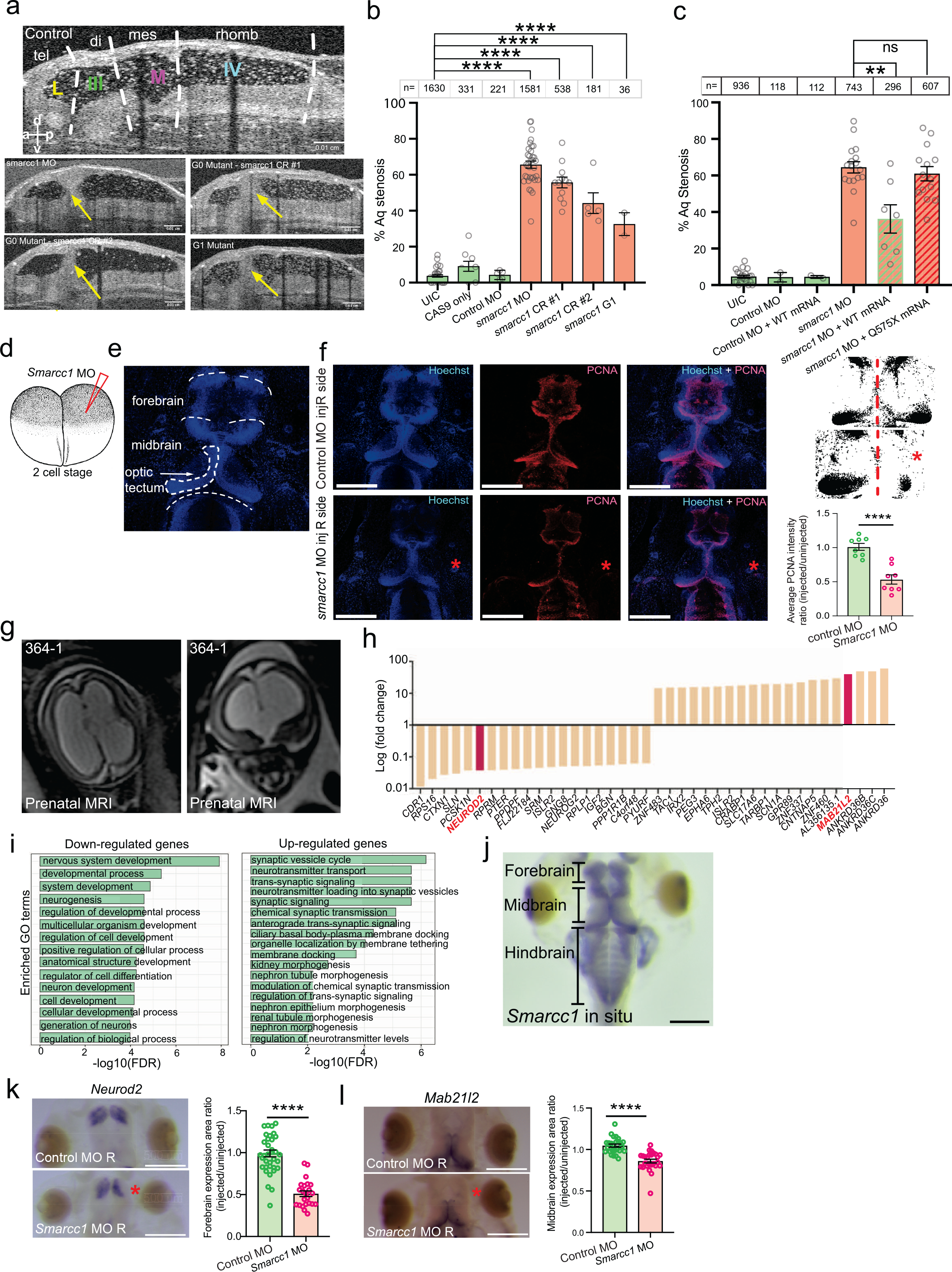
*SMARCC1* mutation causes hydrocephalus by disrupting the epigenetic regulation of gene transcription in periventricular neuroprogenitor cells. A. Mid-sagittal view of the *Xenopus* ventricular system. Dotted white lines indicate boundaries between labeled regions: tel, telencephalon; di, diencephalon; mes, mesencephalon; rhomb, rhombencephalon; L, lateral ventricle; III, third ventricle; M, midbrain ventricle; IV, fourth ventricle. Representative mid-sagittal views for experimental conditions (G0 variant from morpholino oligo, G0 variant from CRISPR #1, G0 variant from CRISPR #2, and G1 variant progeny from *Smarcc1* MO animals) are shown with aqueductal occlusion marked by arrows. B. Quantification of % aqueductal stenosis in uninjected controls (UIC); Cas9 control, and control MO, as well as in the experimental conditions *Smarcc1* MO, *Smarcc1* CRISPR #1, *Smarcc1* CRISPR #2, and *Smarcc1* G1 variant. Data are shown as mean +/- SEM. Open circles indicate the number of experiments, with animal counts indicated above each column. Significance was calculated by one-way ANOVA; **** indicates p ≤ 0.0001. C. Quantification of rescue of aqueductal stenosis phenotype with *Smarcc1* MO + WT mRNA (p = 0.0024) with recapitulation of phenotype by pathogenic mRNA from Q575* variant (p = 0.4888). Data are shown as mean +/- SEM. Open circles indicate number of experiments, with animal counts indicated above each column. Significance was calculated by Mann-Whitney test. D. Schematic of two-cell injection protocol in *X. tropicalis*. E. Labeled representative fluorescence microscopy of WT stage 46 *X. tropicalis* stained with Hoechst. Olfactory bulb, forebrain, midbrain, optic tectum, and cerebellum are indicated. F. Representative immunofluorescence images of right side-injected control MO and right side-injected *Smarcc1* MO stage 46 *X. tropicalis* for PCNA (red) and merged images (with Hoechst, blue). Scale bar represents 500 um. Schematic and chart for quantification of average PCNA intensity ratio for control and *smarcc1* MO injected on the right side with left side un-injected, p ≤ 0.0001 with unpaired t-test. Data are shown as Mean +/- SEM. G. Prenatal ultrasound imaging for patient CHYD364-1 demonstrates ventriculomegaly. H. Median fold-change of top 20 differentially-expressed genes. *NEUROD2* and *MAB21L2* are highlighted. I. GO analysis of CH risk genes, including ranked and selected terms. Significance was calculated by two-sided Fisher’s exact test. Scale bar 500 um. J. Representative photomicrograph of DNA *in situ* hybridization showing *Smarcc1* expression in WT stage 46 *X. tropicalis*. The forebrain, midbrain, and hindbrain are indicated. Scale bar represents 500 um. K. Representative photomicrographs of DNA *in situ* hybridization showing *Neurod2* expression in stage 46 *X. tropicalis*, control and *Smarcc1* MO-injected on the right side, with left side un-injected. Quantification of forebrain expression area is shown, p ≤ 0.0001 with unpaired t-test. Data are shown as Means ± SEM. Scale bar represents 500 um. L. Representative photomicrographs of DNA *in situ* hybridization showing *Mab21l2* expression and quantification as in L. Scale bar represents 500 um.

To begin to elucidate the cellular pathogenesis of *SMARCC1*-mutant hydrocephalus, we leveraged the fate patterning of *Xenopus*, in which embryos at the two-cell stage can be selectively injected with MO on one side of the organism and then compared with the opposite side injected with nonsense MO as an isogenic control (see Methods, Figure 2d). PCNA immunostaining (a marker of cellular proliferation) at stage 46 showed *Smarcc1* variants have significantly fewer PCNA^+^ periventricular cells on the MO-injected side compared to the control side (Figure 2e, 2f, Extended Data Figure 8). This was particularly evident in the midbrain and tectum, structures situated dorsal and ventral to the aqueduct, respectively. The length of the tectum in *Smarcc1* depleted tissue was markedly reduced and its angulation with the anterior portion of the midbrain was altered, indicating significant dysmorphology or atresia. Forebrain thickness was also significantly reduced on the MO-injected versus the control side (Extended Data Figure 8). These data are consistent with cortical and midbrain dysgenesis secondary to the impaired proliferation of *SMARCC1*-mutant NPCs.

To gain insight into the molecular impact of *SMARCC1* mutation in humans, we performed bulk RNA-seq analysis on human frontal and motor-sensory cortex tissue from severely hydrocephalic proband CHYD364-1 (p.Gln575X), who unfortunately underwent fetal demise at PCW19 (Figure 2g). We compared these results to a spatial and developmental time-matched RNA-seq control dataset from the BrainSpan database.^49,50^ Analysis of differentially expressed genes (DEGs) identified several genes with significantly higher or lower expression compared to control samples (Figure 2h, Extended Data Figure 9). Gene Ontology (GO) analysis of significantly down-regulated DEGs showed enrichment in multiple terms related to structural neurodevelopment, including ‘nervous system development,’ ‘neurogenesis,’ and ‘regulation of cell development,’ whereas significantly up-regulated DEGs were enriched for terms related to neural transport and signaling (Figure 2i).

Among the most significant DEGs were *NEUROD2* and, transcription factors with human orthologues in *Xenopus* which have been shown in both mice and *Xenopus* to play critical roles in brain morphogenesis via NPC regulation^50-53,51-54^ (Extended Data Figure 10). Examination of the expression profiles of *NEUROD2* and *MAB21L2* in human prenatal single-cell RNA-sequencing (scRNA-seq) datasets revealed highly enriched expression in intermediate progenitor cells (IPC1), the same cell type with robust *SMARCC1* expression during early brain development (Extended Data Figure 11). Whole-mount *in situ* hybridization showed MO-mediated *Smarcc1* depletion in the two-cell model (see above) caused a significant reduction of *Neurod2* and *Mab21l2*, on the MO-injected versus control side (Figure 2j, 2k, 2l). These results suggest *SMARCC1* mutation in CH may cause cortical and midbrain dysgenesis by altering the expression of key transcription factors, including *Neurod2* and *Mab21l2*, that are involved in the regulation of the growth and proliferation of NPCs.

## DISCUSSION

Our data provide evidence that *SMARCC1* is a *bona fide* CH risk gene and that mutations in *SMARCC1* cause a novel human syndrome characterized by DD, cerebral ventriculomegaly and aqueductal stenosis, and other associated structural brain and cardiac defects. We propose the name “SMARCC1-associated Developmental Dysgenesis Syndrome (SaDDS)” to describe this novel BAFopathy. These results highlight the importance of SMARCC1 and the BAF chromatin remodeling complex in human brain morphogenesis and provide further support for a “neural stem cell” paradigm of human CH.^34,35,55,56^ These data also demonstrate the power of trio-based WES for identifying pathogenic variants in sporadic structural brain disorders, and suggest WES may be a useful prognostic adjunct when evaluating the surgical candidacy of CH patients.

The mutation spectrum of *SMARCC1* involving its BAF complex-interacting SWIRM, Chromo, and Myb-DNA-binding domains suggests that these variants lead to functional impairment of the BAF complex.^32,33,40^ Indeed, NDDs with overlapping clinical phenotypes are associated with pathogenic variants in other subunits of the BAF chromatin remodeling complex. Thus, the *SMARCC1*-associated condition presented here partially overlaps with BAFopathies such as CSS and Nicolaides-Baraitser-like syndrome, characterized by ID/DD and neurobehavioral abnormalities. A defining phenotype of patients with *SMARCC1* variation is cerebral ventriculomegaly and aqueductal stenosis that often requires neurosurgical intervention. This phenotype is likely overrepresented in our patients, insofar as recruitment for our cohort was based on the presence of cerebral ventriculomegaly. However, it is interesting that murine knockout of *Smarca4*, encoding a binding partner of Smarcc1 in the BAF complex, produces severe congenital hydrocephalus with enlargement of the lateral ventricles, aqueductal stenosis, and attenuation of the cerebral cortex. ^57^

Our results suggest incomplete penetrance and variable expressivity for some *SMARCC1* variants, a phenomenon well-recognized for other CH and ASD risk genes^32,58^, as well as for other BAF complex genes.^22,59-61^ Mechanistic drivers of incomplete penetrance and variable expressivity in this specific context remain unclear but may include common genetic or environmental modifiers such as inflammatory or oxidative triggers,^62,63^ as well as stochastic components resulting in mosaicism.^64^ As the paralogous *SMARCC1* and *SMARCC2* gene products form hetero- or homodimers^15^ and share functional scaffolding properties,^28^ upregulation of *SMARCC2* or other BAF members could compensate for the loss of *SMARCC1*, possibly leading to less severe phenotypes. As different *SMARCC1* isoforms are expressed in different tissues and at different developmental stages^24,32,50^, both the spatio-temporal profile and isoform of the protein containing the mutation could affect the phenotype. Further, the wide phenotypic variety and lack of clear genotype-phenotype correlation characterizing conditions associated with BAF-complex mutations suggest possible gene dosage-dependent mechanisms affecting variants of the ATP-dependent chromatin remodeling machinery.^17^

The well-orchestrated spatiotemporal regulation of BAF complex subunit assembly and activity is essential for the development and function of the central nervous system.^26,65,66^ BAF complexes specific to NPCs control cell proliferation, differentiation, and survival through the epigenetic regulation of gene expression that is essential for telencephalon development.^24,27^ We showed *SMARCC1* is highly expressed in fetal human NPCs,^9,29^ and *SMARCC1* mutations dysregulate the expression of genes critical for neurogenesis, including transcription factors *NEUROD2 and MAB21L2*. Our data suggest *SMARCC1* mutations, possibly by altering the epigenetic regulation of gene expression, impair NPC growth and proliferation. Resultant attenuation of neurogenesis and gliogenesis could then lead to non-obstructive (“*ex vacuo*”) ventriculomegaly from a thinned cortical mantle, as has been shown with other CH-associated gene mutations,^55^ and obstructive hydrocephalus secondary to midbrain dysgenesis and aqueductal stenosis. The latter is consistent with the fact that normal cerebral aqueduct development requires the precise regulation of the proliferation and differentiation of NPCs and the development of their associated fiber tracts in the mesencephalon following the prenatal closure of the neural tube.^67-70^

Notably, the BAF complex also modulates the expression of cardiac progenitor cells and regulates cardiac remodeling during development and repair.^11-13,71^ Inactivation of the complex has been implicated in cardiac hypertrophy, shortened or incomplete separation of outflow tracts, and persistent truncus arteriosus in rodents.^11,72^ In particular, a p.Lys615Ile frameshift mutation in *SMARCC1* has been associated with hypoplastic right or left heart syndrome in humans.^73^ Recent studies indicate that congenital heart disease (CHD) patients exhibit increased prevalence of neurodevelopmental disabilities, and CHD patients with neurodevelopmental outcomes have a higher burden of damaging DNMs, particularly in genes important to both heart and brain development. ^74^

Multiple “impaired brain plumbing” mechanisms have been proposed to account for hydrocephalus, including increased CSF secretion, decreased intraventricular CSF transit from cilia dysfunction, and decreased CSF reabsorption associated with elevated venous pressure, arachnoid granulation immaturity, and lymphatic dysplasia. ^75^ However, accumulating genetic data^32,58,76,77^ suggest that impaired neurogenesis rather than overactive CSF accumulation may underlie some forms of CH. Our findings with SMARCC1 support such a “neural stem cell” paradigm of disease.^32,58,76^ In the case of patients with mutations in *TRIM71* (another CH-associated gene), impaired post-transcriptional silencing of RNA targets leads to decreased NPC proliferation at even earlier time points, resulting in severe cortical hypoplasia and secondary ventriculomegaly. ^76^ The opposite phenotype of NPC hyperproliferation due to *PTEN* variant and constitutive activation of the PI3K pathway may also occur. ^78^

The diversity of genetic etiologies and underlying biochemical pathways in CH supports the implementation of routine clinical WES for newly diagnosed patients. Current recommendations for workup of fetal and neonatal ventriculomegaly include rapid commercial microarray testing for known chromosomal and copy-number abnormalities.^79^ However, this strategy does not address CH cases explained by recently detected mutations in many new CH genes^32,33,55^. Application of routine WES or whole genome sequencing could improve the management of children with CH by aiding prognostication and treatment stratification (including when or when not to operate); increasing vigilance for medical screening of mutation-associated conditions (such as cancer surveillance for CH patients with variants in *PIK3CA* or *PTEN*); and providing recurrence rates to increase reproductive confidence.

### Limitations

Our study is limited by the fact our cohort was ascertained on the basis of congenital cerebral ventriculomegaly, including treated hydrocephalus, given previous work in much smaller cohorts suggested a role for SMARCC1in these conditions.^32,33^ Continued identification of mutant patients will expand our knowledge of the SMARCC1 phenotypic spectrum, including those patients without ventriculomegaly.

Additional patients may also permit genotype-phenotype correlations. In addition, brain tissue for bulk RNA-seq was available from only one human subject, since access to *SMARCC1*-mutated fetal brain is such an exceedingly rare event. Using single cell RNA-seq on multiple human subjects and inclusion of biological replicates would be a goal of future investigations. Another limitation is the embryonic lethality phenotype of *Smarcc1* knockout mice,^29,30^ which necessitated our use of the non-mammalian *Xenopus* model for disease modeling and mechanistic study.

## Conclusions

*SMARCC1* is a *bona fide* CH risk gene. Pathogenic *SMARCC1* variants cause a novel human BAFopathy characterized by DD, cerebral ventriculomegaly, and a variety of structural brain or cardiac defects. These data underscore the importance of SMARCC1 and the BAF chromatin remodeling complex for human brain morphogenesis and support a neural stem cell paradigm of human CH pathogenesis. These results highlight the power of trio-based WES for identifying risk genes for congenital structural brain disorders and suggest WES may be a valuable adjunct in the management of patients with CH.

## METHODS

We confirm that our research complies with all relevant ethical regulations as approved by the Yale University Human Investigation Committee, the Yale University Institutional Animal Care and Use Committee, the Washington University in St. Louis Institutional Animal Care and Use Committee, and the Massachusetts General Hospital Institutional Animal Care and Use Committee.

### Statistics and Reproducibility

No power analysis was performed to predetermine sample size, as our sample sizes are similar to those reported in previous publications^32,33,80^. Randomization was not relevant to this study as controls and *X. tropicalis* knockdowns did not receive different treatments and human studies were descriptive studies. All experiments were performed and analyzed in a blinded manner. No data were excluded from the analyses. Wilcoxon Rank-Sum test was used in differential gene expression analysis, as described in Methods. Mann-Whitney test was used to analyze experimental data in Figure 2c. Elsewhere, data distribution was assumed to be normal, but this was not formally tested.

### Patient cohort

All study procedures and protocols were guided by and in compliance with ‘Human Investigation Committee and Human Research Protection Program at Yale University and the Massachusetts General Hospital. All participants provided written, informed consent to participate. For patients from the clinical laboratory GeneDx, denoted CHYDX, written informed consent for genetic testing was obtained from the guardians of all pediatric individuals undergoing testing. The Western institutional review board waived authorization for the use of de-identified aggregate data for the purposes of this study. Criteria for inclusion into the study was congenital or primary cerebral ventriculomegaly, including congenital hydrocephalus. Patients and participating family members provided buccal swab samples (Isohelix SK-2S DNA buccal swab kits), medical records, neuroimaging studies, operative reports, and phenotype data when available. Human phenotype ontology terms were used to aggregate relevant pediatric patients in the GeneDx database. The comparison control cohort consisted of 1,798 unaffected siblings of people diagnosed with autism spectrum disorder (ASD) and unaffected parents sourced from the Simons simplex consortium (SSC)^40 81^. Only the unaffected siblings and parents, as designated by SSC, were included in the analysis, and served as controls for this study. Permission to access the genomic data in the SSC on the National Institute of Mental Health Data Repository was obtained. Written and informed consent for all participants was provided by the Simons Foundation Autism Research Initiative.

### Kinship analysis

Pedigree information and relationships between proband and parents was confirmed using pairwise PLINK identity-by-descent (IBD) calculation.^82^ The IBD sharing between the probands and parents in all trios was between 45% and 55%. Pairwise individual relatedness was calculated using KING^78 83^. The ethnicity of each patient from the Yale cohort was determined by single-nucleotide polymorphisms in cases, controls, and HapMap samples using EIGENSTRAT, as previously described.^84^ For the GeneDx cohort, kinship analysis was performed using an internally developed KNN/PCA pipeline.

### WES and variant calling

Patient genomic DNA samples derived from saliva or blood were applied for exon capture using Roche SeqCap EZ MedExome Target Enrichment kit or IDT xGen target capture followed by 101 or 148 base-paired-end sequencing on Illumina platforms as described previously^32,33^. BWA-MEM was applied to align sequence reads to the human reference genome GRCh37/hg19. Single-nucleotide variants and small indels were called using a combination of GATK HaplotypeCaller and Freebayes^85,86^ and annotated using ANNOVAR^81 39^. Allele frequencies were annotated in the Exome Aggregation Consortium, GnomAD (v.2.1.1) and Bravo databases.^87^ Variant filtration and analysis were conducted following GATK best practices and consensus workflows.^88^ MetaSVM and MPC algorithms were used to predict the deleteriousness of missense variants (D-mis, defined as MetaSVM-deleterious or MPC-score ≥2). ^89^ Inferred loss-of-function (LoF) variants consisted of stop-gain, stop-loss, frameshift insertions/deletions, canonical splice site and start-loss. LoF and D-Mis variants were considered ‘damaging’. Analyses were conducted separately for each class of variant – *de novo* variants (DNVs), homozygous recessive variants, and rare, heterozygous dominant variants – following previously established analytical methodologies^33,88^. Firstly, DNVs from the Yale cohort were called from all CH parent-offspring trios using the established TrioDeNovo pipeline^90,91^. GeneDx DNVs were called as previously defined.^92^ Candidate DNVs for all samples were further filtered based on whether the variants were called in the exonic or splice-site regions, the variant read depth (DP) was at least ten in the proband as well as both parents, and the global minor allele frequency was less than or equal to 4 × 10^−4^ in the Exome Aggregation Consortium database. Samples from the Yale cohort were subsequently filtered based on the following criteria: (i) the proband’s alternative read depth was greater than or equal to five; (ii) proband alternative allele ratio greater than or equal to 28% if having less than ten alternative reads, or less than or equal to 20% if having greater than or equal to ten alternative reads; (iii) the alternative allele ratio in both parents less than or equal to 3.5%. Samples from the GeneDx cohort were additionally filtered based on the following criteria: (i) Genotype quality (GQ) > 40 for all family members; (ii) Variant quality score log odds (VQSLOD) > -10; (iii) Phred-scaled p-value (Fishers exact test; FS) <30; (iv) Proband alternate allele count >4; (v) Proband alternate allele ratio > 0.1; (vi) Proband alternate allele ratio >0.15 if REF and ALT calls are of equal length; (vii) Proband alternate allele ratio >0.25 if REF and ALT calls are of unequal length; (viii) Proband alternate allele ratio < 0.9 in proband if DNV is autosomal; (ix) DNV must be < 100 bps in size for both the REF and ALT calls; (x) If the VQSLOD <7 and the alternate allele ratio in the proband <0.3, the variant was omitted. (xi) DNVs were omitted if they existed in more than 2 unrelated probands. After filtering as above, *in silico* visualization was performed, applying in-house software to manually inspect each variant for false-positive calls. Variants found to be false-positive upon manual inspection were removed. *SMARCC1* variant annotations were then confirmed through manual cross-reference in the UCSC Genome Browser.^38,39^ Reported variants passing these filters and manual inspection in SMARCC1 were further confirmed by Sanger sequencing.

#### Developmental human brain scRNA-seq dataset analysis

As described previously^24^, the preprocessing and clustering analysis for ScRNA developmental human brain dataset was completed using Seurat.^93^ Briefly, cells with fewer than 1000 genes/cell were removed, as were cells with greater than 10% of their individual transcriptome represented in either mitochondrial or ribosomal transcripts. Only genes expressed in at least 30 cells were carried forward in the analysis. The raw counts were normalized and log2 transformed by first calculating ‘size factors’ that represented the extent to which counts should be scaled in each library. Highly variable genes were detected using the proposed workflow of and were subsequently used for unsupervised dimensionality reduction techniques and principal component analysis. UMAP coordinates were calculated using standard Seurat workflow, and clusters were assigned to cells based on previous analysis via a hybrid method using Louvain clustering and WGCNA.^24^ Non-parameteric Wilcoxon rank sum test was used to identify differentially expressed markers across time points, areas and laminar zones by running FindAllMarkers. Heatmap expression values were calculated using AverageExpression function and visualization of the heatmaps were created using pheatmap package.

### Cell type enrichment

Cell type enrichment for the expression of SMARCC1 was tested in scRNA-seq datasets of prenatal human brain using in-house custom-made script in R studio^49,94^. Enrichment for each cell type was tested using hypergeometric test, where a gene list was significantly enriched in a cell type if the adjusted *p* value was less than 0.05. Average expression was shown using the DotPlot function from the Seurat package. For control comparison, we used frontal neocortex layer-specific data from the BrainSpan database matched by developmental age. ^94^ Batch correction was applied by quantile normalization in the limma package.^95^ Median fold changes of gene expression were used to rank the genes. Only protein-coding genes were used for analysis.

### *SMARCC1* expression in PsychENCODE bulk RNA sequencing

To examine the expression pattern of SMARCC1 during human brain development, we extracted RPKM expression from the PsychENCODE bulk tissue RNA sequencing dataset.^49^ Gene expression was scaled, centered and average values calculated across developmental periods. The expression distributions were visualized in a violin plot.

### Gene Ontology (GO) enrichment analysis

To test for functional enrichment for all modules and the respective genes, we performed gene ontology enrichment analysis (GOEA) for using the GO set of biological processes. Gene set ‘GO. v5.2.symbols_mouse.gmt’ was obtained from the Molecular Signatures Database. The compareCluster and enrichGo functions from the R package ClusterProfiler (version 3.12.0) were used to determine significant enrichment (*q* < 0.05) of biological processes. All present genes were used as background (universe). To focus only on neurological gene sets, GO term gene sets were selected for terms including the term ‘neuro’, ‘neural’ and ‘nerv’. Network visualization was performed using the cnetplot function from the R package ClusterProfiler.

### *Xenopus* husbandry

*Xenopus tropicalis* were raised and cared for in our aquatics facility according to protocols approved by the Yale University Institutional Animal Care and Use Committee. Embryos were staged according to Nieuwkoop and Faber^90 96^.

### sgRNA and RNA production

CRISPR: Two non-overlapping CRISPR sgRNAs were designed on crisprscan.org for the *Xenopus tropicalis smarcc1* gene (Xenbase genome v9.1) and produced using an EnGen sgRNA Synthesis Kit (NEB # E3322). Target sites are located in exon #10 (CRISPR #1: 5’-AGGCTGTGCGCAGTCCCGAGAGG-3’), and exon 1 (CRISPR #2: 5’-CGGCCGGGAAGAGCCCCGCAGGG-3’. CRISPR indels were verified by performing Sanger sequencing on PCR products using genomic DNA from stage 46 embryos. Genomic DNA was extracted from individual anesthetized embryos at phenotypic stage 46 by 10 minute incubation in 50ul of 50mM NaOH at 95°C followed by neutralization with 20ul of 1M Tris pH 7.4. PCR was performed with either Phusion High-Fidelity DNA Polymerase (NEB #M0530) (CRISPR #1), or Platinum SuperFi II Green PCR Master Mix (Thermo Fisher Scientific #12369050) (CRISPR #2) using primers around the CRISPR cut site (CRISPR #1: 5’-ACATTGGTCCCTGTGCTTTT-3’ and 5’-TTCAAGTCCTCGTCTGTTTGG-3’, CRISPR #2: 5’-AACGGCAGCAATAACGGAGA-3’ and 5’-AGATACATGTCCCCTCCGCA-3’). PCR sequences were analyzed for indels using the online Inference of CRISPR Edits (ICE) tool (Synthego).

Human mRNA: Human SMARCC1 mRNA was produced by cloning a full length insert (sequence ID NM_003074.4) into a pCS DEST expression plasmid backbone using Gateway recombination techniques. mRNA was synthesized using a mMESSAGE mMACHINE SP6 Transcription Kit (Thermo Fisher Scientific #AM1340). The patient variant 1723T>C (Q575X) was produced using inverse PCR. Overlapping primers [forward and reverse] were designed with the base change located in the middle, with 14 bases on either side of the variant. Long range PCR was performed with wild-type plasmid template using Platinum Taq DNA Polymerase High Fidelity (Thermo Fisher Scientific # 11304011). DpnI digestion removed methylated template DNA from nonmethylated PCR product, which was then used to transform Ca^2+^-competent E. Coli. Individual clones were grown up, and plasmid DNA was extracted and sequenced to verify the presence in the insert of only the desired mutation. The insert was cloned into a pCS DEST expression vector and mRNA was synthesized as above.

### Microinjection, gene expression, knockdown and overexpression

*Xenopus tropicalis* embryos were microinjected using standard protocols.^97-99^ Fertilized eggs were injected at either 1 cell stage with a 2 nl volume, or into 1 of 2 cells at 2 cell stage with a 1 nl volume. Injection mixes for expression knockdown using morpholino oligo (MO) consisted of the fluorescent tracer Dextran Alexa Fluor 488 (Thermo Fisher Scientific #D22910) along with MO targeting the start site of *Xenopus tropicalis smarcc1* (gene model XM_002942718.5) (5’-CCTTTGTTTCATGGCTGCTACTCCC-3’, Gene Tools) at a concentration leading to a dose of 0.5-1.0ng per embryo. A standard control MO (5′-CCTCTTACCTCAGTTACAATTTATA-3′, Gene Tools) was also used at the same dose. We also used CRISPR/Cas9-mediated gene editing for expression knockdown as previously described^91 100^. CRISPR injection mixes consisted of fluorescent tracer, and the following components at a concentration leading to the listed dose per embryo: 1.6 ng Cas9 (CP03, PNA Bio), 400pg sgRNA. 500 pg mRNA encoding human wild type or patient variant protein was injected into each embryo to rescue MO knockdown. Injections were verified using a Zeiss SteREO Lumar.V12 microscope to visualize fluorescence at stages 18-46 in the entire embryo (1 cell injection), or on only the right or left side (2 cell injection).

### Optical coherence tomography imaging

A Thorlabs Ganymede II HR OCT imaging system using ThorImage OCT version 5.0.1.0 software was used to obtain 2D cross-sectional images and movies. Imaging was obtained as previously described.^98^

### Western blotting

Whole cell lysate was extracted from pooled embryos in RIPA lysis buffer (Millipore #20-188) supplemented with Halt protease and phosphatase inhibitor cocktail (Thermo Fisher Scientific #78440). Samples were run on Bolt 4-12% Bis-Tris Plus gels (Thermo Fisher Scientific) then transferred to PVDF membranes. Immunobloting was performed using standard methods. Polyclonal rabbit anti-SMARCC1 antibody (Thermo Fisher Scientific #PA5-96513) was used at 1:1000, and mouse anti-b-Actin (C4) HRP (Santa Cruz Biotechnology #sc47778) was used at 1:100,000 as a loading control.

### Whole mount *in situ* hybridization

Whole mount in situ hybridization (WISH) was performed as previously described^92 101^. Briefly, *Xenopus tropicalis* embryos were fixed in 4% paraformaldehyde in 2mM EGTA, then dehydrated through methanol washes and stored at -20°C. Embryos were rehydrated through washes in PBS + 0.1% tween-20, then incubated in 4% hydrogen peroxide in PBS + 0.1% Tween-20 to remove pigment. After post-fixing in 4% paraformaldehyde in 2mM EGTA, embryos were hybridized overnight at 60°C with RNA probes. The Digoxigenin-11-UTP (Sigma #11209256910) labeled RNA probes were produced with full length insert containing expression plasmids using either HiScribe T7 (antisense), or HiScribe SP6 (sense) RNA Synthesis Kits (NEB) according to the manufacturers’ instructions.

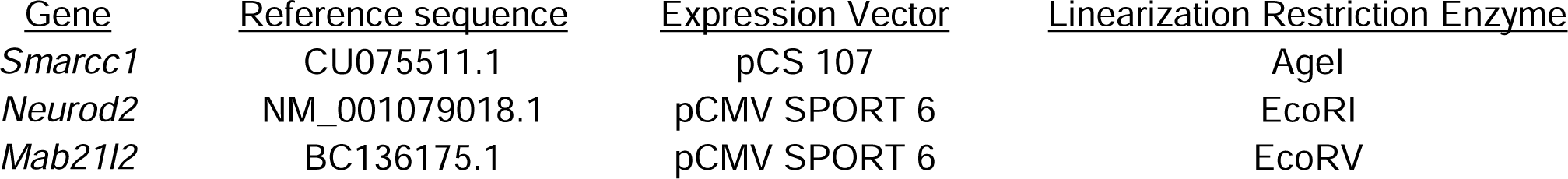

After overnight hybridization, embryos were washed, blocked, then incubated overnight in Anti-Digoxigenin-AP, Fab fragments (Sigma #11093274910). After washes, embryos were incubated in BM Purple (Sigma # 11442074001) until signal was fully visible, then fixed in 4% paraformaldehyde + 0.1% glutaraldehyde in 2mM EGTA.

### Immunohistochemistry

Uninjected control, 1 of 2 cell control MO-, or 1 of 2 cell smarcc1 MO-injected stage 47 embryos were anesthetized, then fixed in 4% paraformaldehyde in PBS for 1 hour at room temperature. After washes in PBS, tails, guts, and ventral structures of the head including lower jaws and facial cartilage were removed. Pigment was bleached from samples by incubation in 5% formamide + 1.2% H_2_O_2_ in PBS while exposed to light. Samples were then washed with PBS + 0.1% Triton X-100 (PTr), blocked in 10% CAS-Block (Thermo Fisher Scientific #008120) in PTr, then incubated in mouse monoclonal anti-PCNA (PC10) (Thermo Fisher Scientific #13-3900) diluted 1:200 in 100% CAS-Block overnight at 4°C. After extensive washes in PTr, samples were blocked in 10% CAS-Block, then incubated overnight at 4°C in Texas Red-conjugated goat anti-mouse IgG (Thermo Fisher Scientific #T-6390) plus Hoechst 33342 (Thermo Fisher Scientific #H3570), respectively diluted 1:200 and 1:5000 in 100% CAS-Block. Samples washed first in PTr, then in PBS, were mounted between two coverslips in ProLong Gold Antifade Mountant (Thermo Fisher Scientific #P36934). Images were obtained using a Zeiss LSM 880 airyscan confocal microscope.

#### RNA sequencing analysis

Sequenced reads were aligned and quantified using STAR: ultrafast universal RNA-seq aligner (version 2.7.3a) and the murine reference genome, GRCm38p5, from the Genome Reference Consortium. Raw counts were imported using the DESeqDataSetFromHTSeqCount function from DESeq2 (version 1.26.0) and rlog-transformed according to the DESeq2 pipeline. DESeq2 was used for calculation of normalized counts for each transcript using default parameters. All normalized transcripts with maximum overall row mean < 20 were excluded, resulting in 13,284 present protein-coding transcripts. Undesired or hidden causes of variation, such as batch and preparation date, were removed using the sva package. The normalized rlog-transformed expression data were adjusted with four surrogate variables identified by sva using the function removeBatchEffect from the limma package. To determine gene clusters, CoCena (Construction of Co-expression network analysis) was calculated based on Pearson correlation on all present genes. Pearson correlation was performed using the R package Hmisc (version 4.1-1). To increase data quality, only significant (*P* < 0.05) correlation values were kept. A Pearson correlation coefficient cutoff of 0.803 (present genes; 10,260 nodes and 69,986 edges) was chosen, resulting in networks following the power-law distribution of *r*2 = 0.934 (scale-free topology). Unbiased clustering was performed using the ‘leiden modularity’ algorithm in igraph (version 1.2.1). Clustering was repeated 100 times. Genes assigned to more than ten different clusters received no cluster assignment. The mean group fold change expression for each cluster and condition is visualized in the Cluster/Condition heat map. Clusters smaller than 40 genes are not shown.

## Data Availability

All data produced in the present study will be made available upon reasonable request from Dr. Kahle at.

## Author affiliations

Department of Neurosurgery, Yale University, New Haven, Connecticut (Singh, Allington, Kiziltug, Mekbib, Shohfi, Duy, DeSpenza, Furey, Reeves, Smith, Allocco); Department of Neurosurgery, Massachusetts General Hospital, Harvard Medical School, Boston, Massachusetts (Singh, Allington, Kiziltug, Mekbib, Shohfi, Duy, DeSpenza, Reeves, Smith, Kahle); Department of Pediatrics, Yale University, New Haven, Connecticut (Viviano, Sestan, Deniz); Department of Genetics, Yale University, New Haven, Connecticut (Viviano, Allington, Ma, Nelson-Williams, Sestan, Deniz); GeneDx, Gaithersburg, Maryland (McGee, Kruszka); Department of Neuroscience, Yale University, New Haven, Connecticut (Duy, Despenza, Ma); Waisman Center, University of Wisconsin-Madison, Madison, Wisconsin (Sousa); Department of Pharmaceutical and Biological Chemistry, University College London School of Pharmacy, London, United Kingdom (Haider, Rizvi); UCL Centre for Advanced Research Computing, University College London, United Kingdom (Haider); Broad Institute of MIT and Harvard, Cambridge, Masachusetts (Alper, Kahle); Division of Nephrology and Vascular Biology Research Center, Beth Israel Deaconess Medical Center, Boston, Massachusetts (Alper); Department of Medicine, Harvard Medical School, Boston, Massachusetts (Alper); Autism & Developmental Medicine Institute, Geisinger, Danville, Pennsylvania (Shimelis, Walsh, Moreno-De-Luca); Laboratory of Human Genetics and Genomics, The Rockefeller University, New York, New York (Lifton); Department of Radiology, Diagnostic Medicine Institute, Geisinger, Danville, Pennsylvania (Moreno-De-Luca); Departments of Genetics and Pediatrics, Washington University School of Medicine, St Louis, Missouri (Jin); Division of Genetics and Genomics, Boston Children’s Hospital, Boston, Massachusetts (Kahle).

## Author contributions

Dr. Kahle and Dr. Deniz had full access to all the data in the study and take responsibility for the integrity of the data and the accuracy of the data analysis.

Concept and design: Kahle.

Acquisition, analysis, or interpretation of data: Singh, Viviano, Allington, Kiziltug, McGee, Mekbib, Duy, Shohfi, Ma, Despenza, Furey, Reeves, Smith, Ma, Sousa, Cherskov, Allocco, Nelson-Williams, Haider, Rizvi, Sestan, Walsh, Shimelis, Moreno-De-Luca, Jin, Deniz, Kahle.

Drafting of the manuscript: Singh, Allington, Sestan, Lifton, Kruszka, Jin, Deniz, Kahle.

Critical revision of the manuscript for important intellectual content: Singh, Viviano, Allington, Sestan, Kruszka, Duy, Ma, Alper, Moreno-De-Luca, Jin, Deniz, Kahle

## Data access, responsibility, and analysis

Dr. Kahle and Dr. Deniz had full access to all the data in the study and takes responsibility for the integrity of the data and the accuracy of the data analysis.

## Data sharing statement

Data will be made available upon reasonable request from Dr. Kahle at.

## Additional contributions

We are grateful to the patients and families who participated in this research.

