## Extended Data Figure 1 for "A novel *SMARCC1*-mutant BAFopathy implicates epigenetic dysregulation of neural progenitors in hydrocephalus"

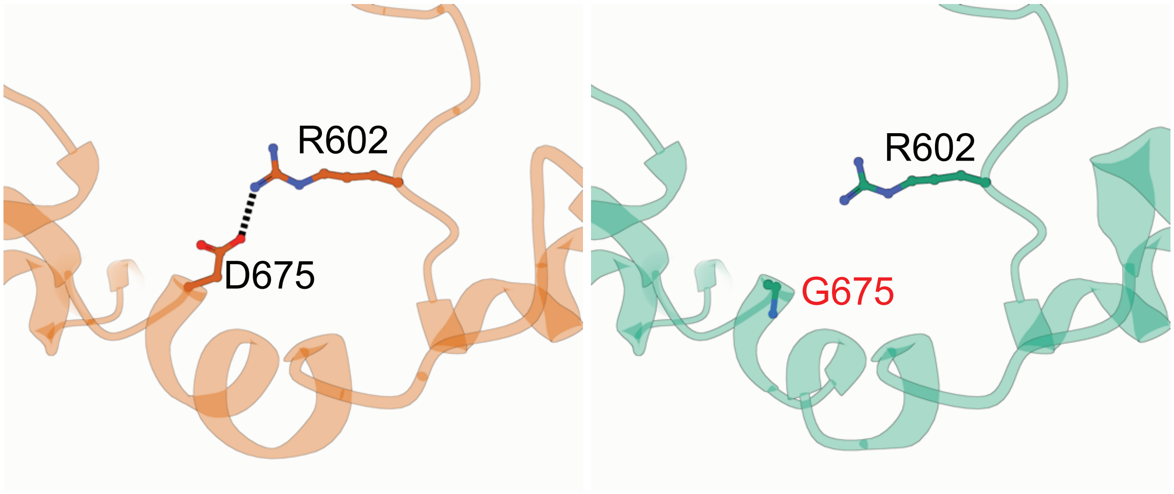


**Extended Data Figure 1.**

p.Asp675Gly was predicted to be detrimental to SMARCC1 structure and function by Alpha-Fold biophysical modelling.

Structural protein modeling predicts that p.Asp675Gly alters a conserved residue in the Myb domain resulting in loss of an ion pair interaction with p.Arg602, with a predicted ΔG 0.73 kcal/mol.
