## Extended Data Figure 2 for "A novel *SMARCC1*-mutant BAFopathy implicates epigenetic dysregulation of neural progenitors in hydrocephalus"

a

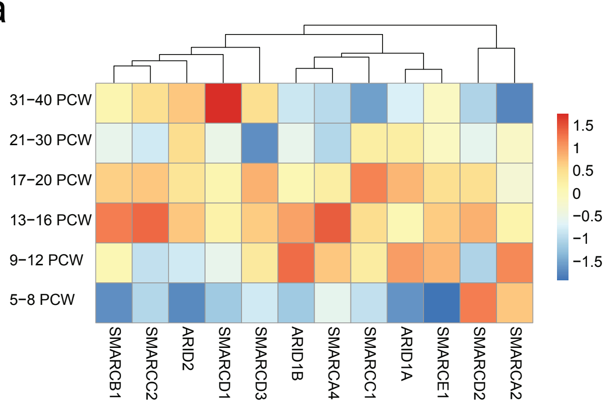


b

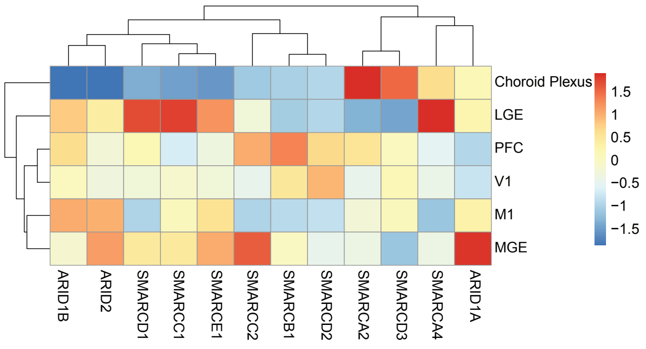


**Extended Data Figure 2.**

SMARCC1 is highly expressed in the LGE between PCW 17-20

A. Analyzed transcriptomic dataset[24] showing heatmap of gene expression levels for SMARCC1 and other BAF complex genes across different developmental timepoints. Vertical axis shows timepoints in post-conception weeks.

B. Analyzed transcriptomic dataset[24] showing heatmap of gene expression levels for SMARCC1 and other BAF complex genes across different brain regions. V1, primary visual cortex; PFC, prefrontal cortex; MGE, medial ganglionic eminence; LGE, lateral ganglionic eminence; M1, primary motor cortex
