## Extended Data Figure 3 for "A novel *SMARCC1*-mutant BAFopathy implicates epigenetic dysregulation of neural progenitors in hydrocephalus"

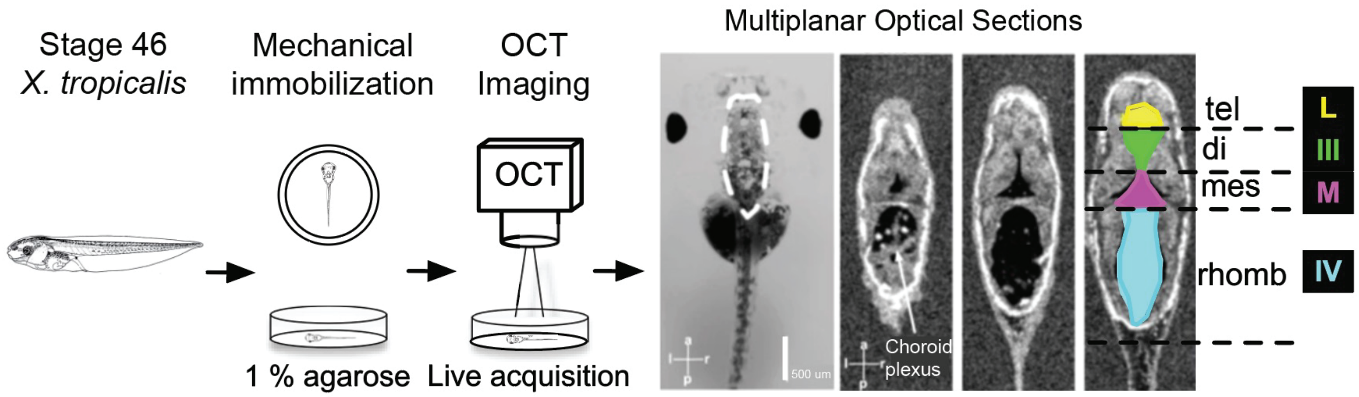


**Extended Data Figure 3.**
Schematic showing Stage 46 tadpole embedded in 1% low-melt agarose and imaged in sections with optical coherence tomography.

The far left photograph shows a stage 46 tadpole under light microscopy. Transverse sections through the ventricular system along the anterior-posterior axis are labeled: choroid plexus; tel, telencephalon; di, diencephalon; mes, mesencephalon; rhomb, rhombencephalon; L, lateral ventricle; III, third ventricle; M, midbrain ventricle; IV fourth ventricle.
