## Extended Data Figure 4 for "A novel *SMARCC1*-mutant BAFopathy implicates epigenetic dysregulation of neural progenitors in hydrocephalus"

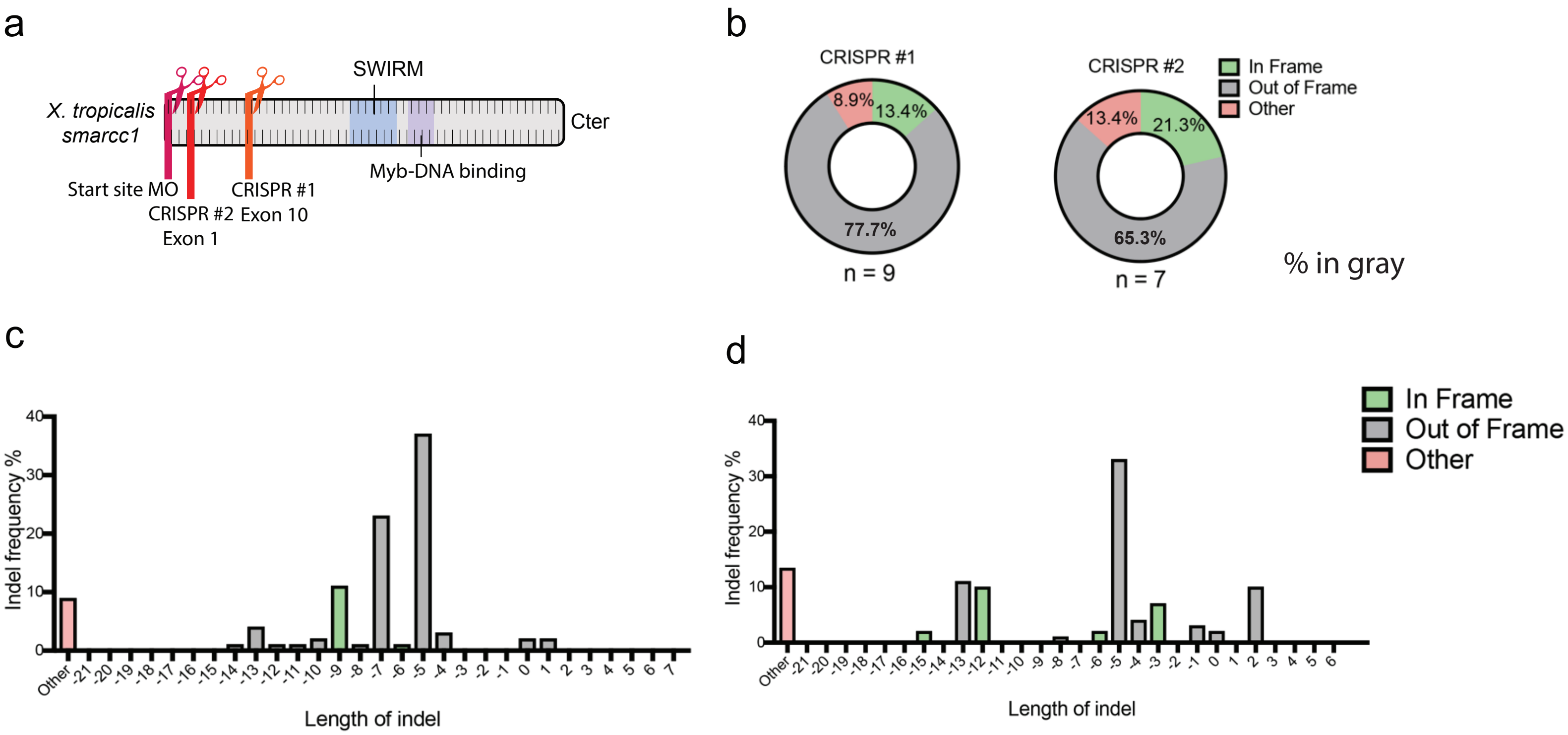


**Extended Data Figure 4.**
Schema of Smarcc1 morphants and knockdowns in *X. tropicalis* with ICE experiments for CRISPR validation.

A. Schematic showing binding and cleavage sites for smarcc1 morpholino oligo, CRISPR #1 in Exon 10 and CRISPR #2 in Exon 1 in relation to the SWIRM domain and Myb-DNA binding domains of Smarcc1.
B. Inference of CRISPR edits (ICE) analysis showing proportion of in-frame and out-of-frame deletion for both constructs.
C. Frequency of in-frame and out-of-frame deletions by length of indel for CRISPR #1.
D. Frequency of in-frame and out-of-frame deletions by length of indel for CRISPR #2.
