## Extended Data Figure 5 for "A novel *SMARCC1*-mutant BAFopathy implicates epigenetic dysregulation of neural progenitors in hydrocephalus"

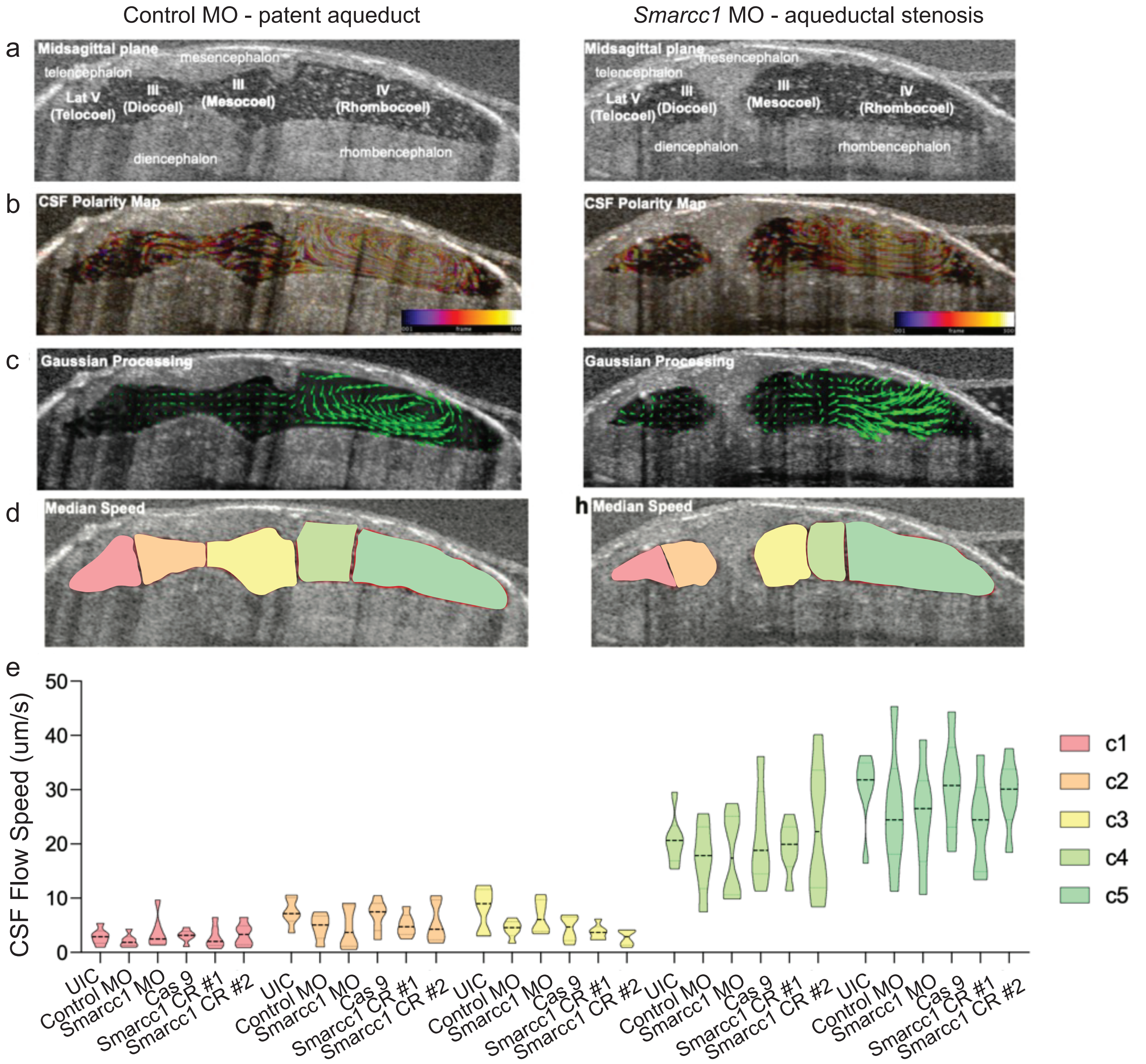


**Extended Data Figure 5.**

Local peri-ventricular wall CSF flow velocities segmented by ventricular region do not differ between control and *Smarcc1* MO, indicating no defect in cilia-driven microcurrents.

A. Midsagittal optical coherence tomography section of stage 46 *X. tropicalis* tadpole. Control morphant and *Smarcc1* are shown. Labelled structures: telencephalon, diencephalon, mesencephalon, rhombencephalon; Lat V, lateral ventricle; III, third ventricle; IV, fourth ventricle.

B. CSF polarity map based on temporary color coded frames delineates particle flow directions.

C. Gaussian processing regression enables detection of flow vectors for quantitation. (see Methods).

D. Violin plot showing local peri-ventricular wall CSF flow speed compared across experimental conditions and different segmented regions. Two-way ANOVA with p-value < 0.0001 for comparison by region, p-value 0.2161 for comparison by experimental condition, and p-value 0.9666 for interaction.
