## Extended Data Figure 6 for "A novel *SMARCC1*-mutant BAFopathy implicates epigenetic dysregulation of neural progenitors in hydrocephalus"

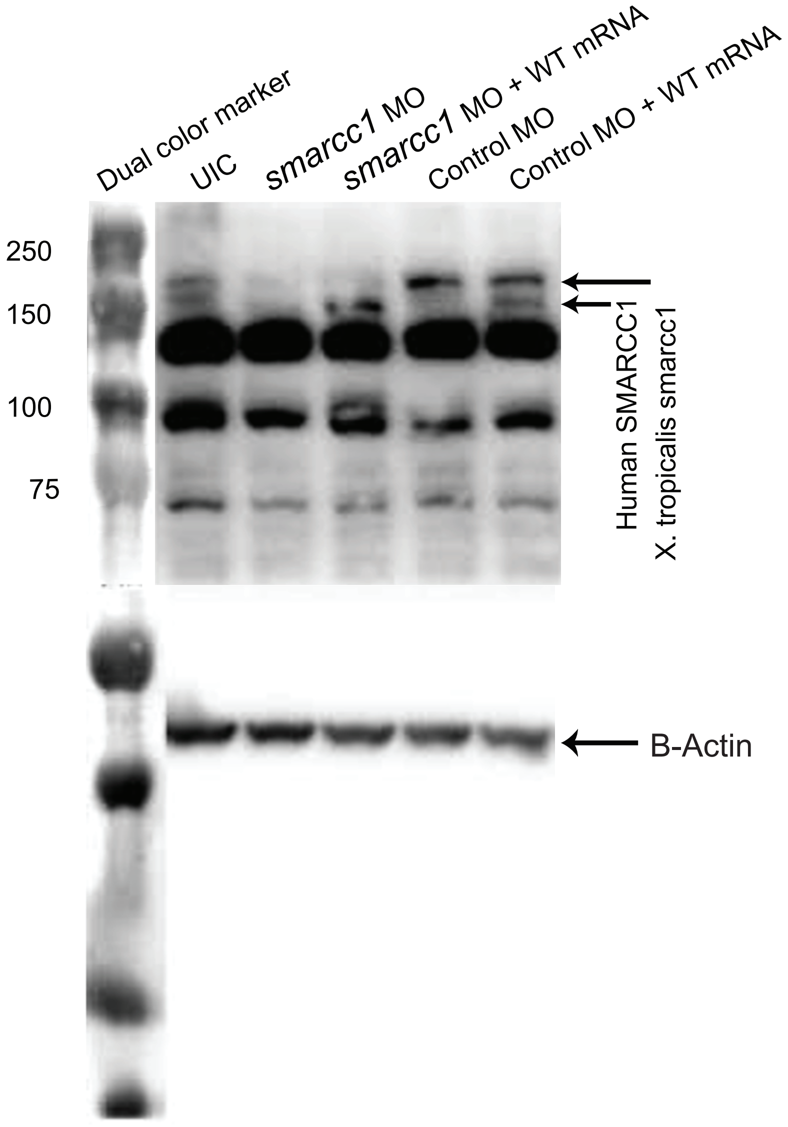


**Extended Data Figure 6.**
Representative western blot shows depletion of Smarcc1 after knockdown in *X. tropicalis*, as well as repletion with the human homolog SMARCC1. A probe for β-Actin was used as a loading control.
