## Extended Data Figure 7 for "A novel *SMARCC1*-mutant BAFopathy implicates epigenetic dysregulation of neural progenitors in hydrocephalus"

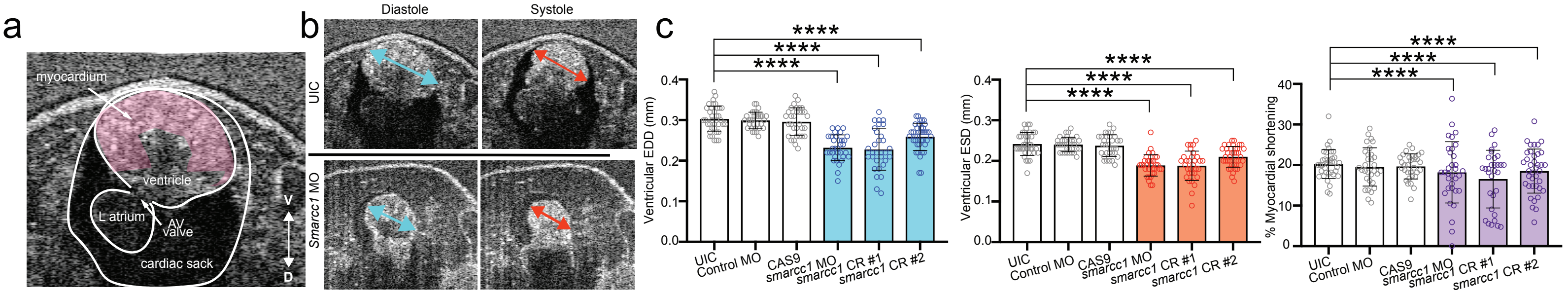


**Extended Data Figure 7.**
Smarcc1 knockdown causes significantly reduced end diastolic diameter (EDD), end systolic diameter (ESD), and myocardial shortening fraction compared to controls.

A. Representative *X. tropicalis* cardiac optical coherence tomography image on ventral-dorsal axis, the ventral three chamber view is shown. Labeled structures are myocardium, ventricle, L atrium, AV valve, and cardiac sack.
B. Representative cardiac measurements by OCT shown for control MO and smarcc1 MO.
C. Quantification of EDD, ESD, and myocardial shortening fraction in UIC, Cas9 control, and control MO, as well as experimental conditions smarcc1 MO, smarcc1 CRISPR #1, smarcc1 CRISPR #2.
Data are shown as Mean +/- SEM.
Significance was calculated by One-way ANOVA using GraphPad Prism where $p \leq 0.0001$ for ****.
