## Extended Data Figure 8 for "A novel *SMARCC1*-mutant BAFopathy implicates epigenetic dysregulation of neural progenitors in hydrocephalus"

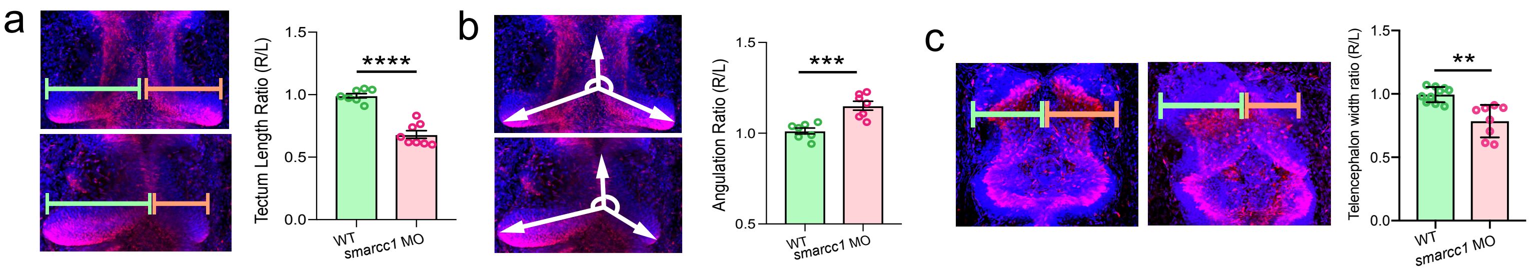


**Extended Data Figure 8.**
PCNA staining in two cell-stage injected embryos reveals dysplastic changes in midbrain and forebrain.

A. Schematic and quantification of optic tectum length ratio for WT control and Smarcc1 MO injected on the right side with left side un-injected, $p \leq 0.0001$ with unpaired t-test. Data are shown as Mean +/- SEM.
B. Schematic and quantification of optic tectum angulation ratio for WT control and Smarcc1 MO injected on the right side with left side un-injected, $p = 0.0008$ with unpaired t-test. Data are shown as Mean +/- SEM.
C. Schematic and quantification of telencephalon width ratio for WT control and Smarcc1 MO injected on the right side with left side un-injected, $p = 0.0019$ with unpaired t-test. Data are shown as Mean +/- SEM.
