## Extended Data Figure 9 for "A novel *SMARCC1*-mutant BAFopathy implicates epigenetic dysregulation of neural progenitors in hydrocephalus"

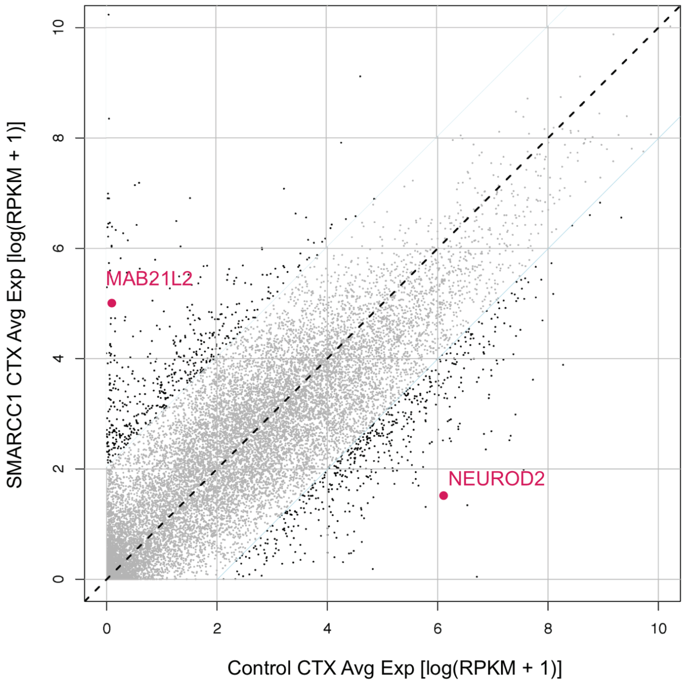


**Extended Data Figure 9.**

Dot plot showing genes differentially expressed by SMARCC1 variant and control samples

Each dot represents a gene. X and y axes respectively represent average gene expression in control and mutant samples. Genes with log-fold change >5 between samples are in black; others are in grey. MAB21L2 and NEUROD2 are highlighted with text.
