## Extended Data Figure 10 for "A novel *SMARCC1*-mutant BAFopathy implicates epigenetic dysregulation of neural progenitors in hydrocephalus"

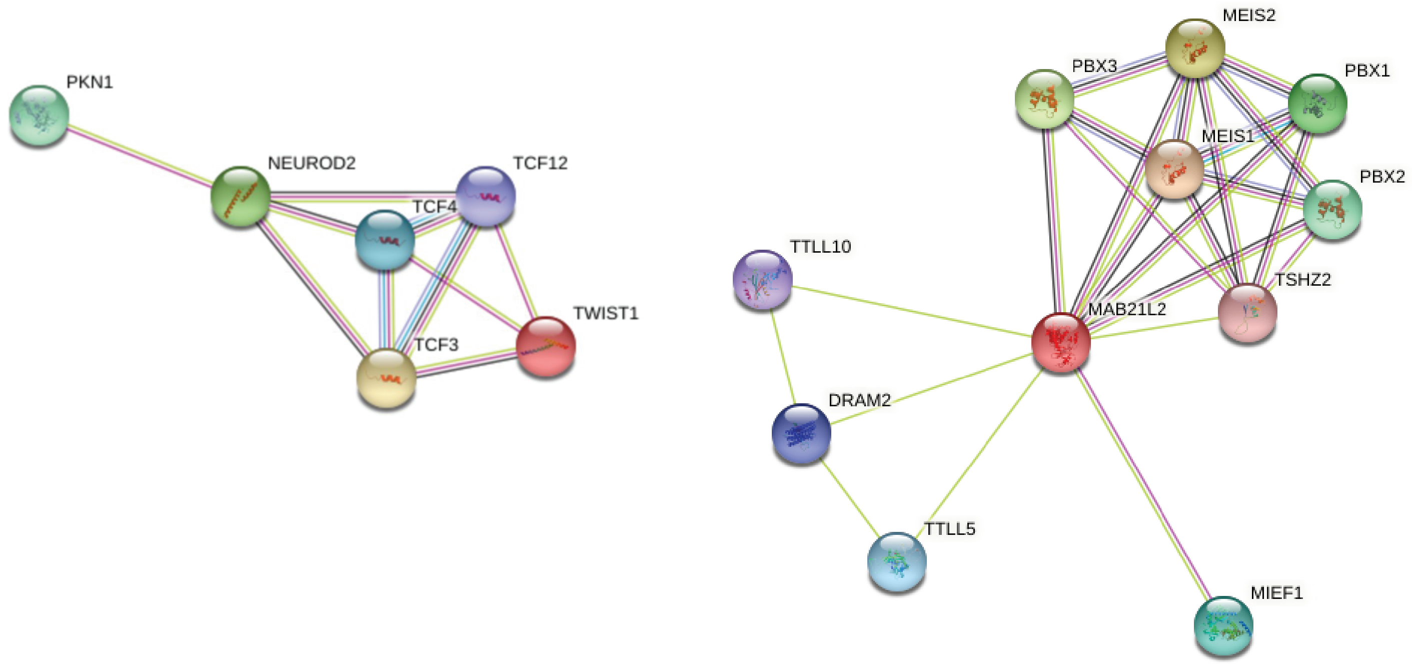


**Extended Data Figure 10.**

Protein network interactions for *NEUROD2* (left) and *MAB21l2* (right). Functional enrichment analysis from the STRING database.
