## Extended Data Figure 11 for "A novel *SMARCC1*-mutant BAFopathy implicates epigenetic dysregulation of neural progenitors in hydrocephalus"

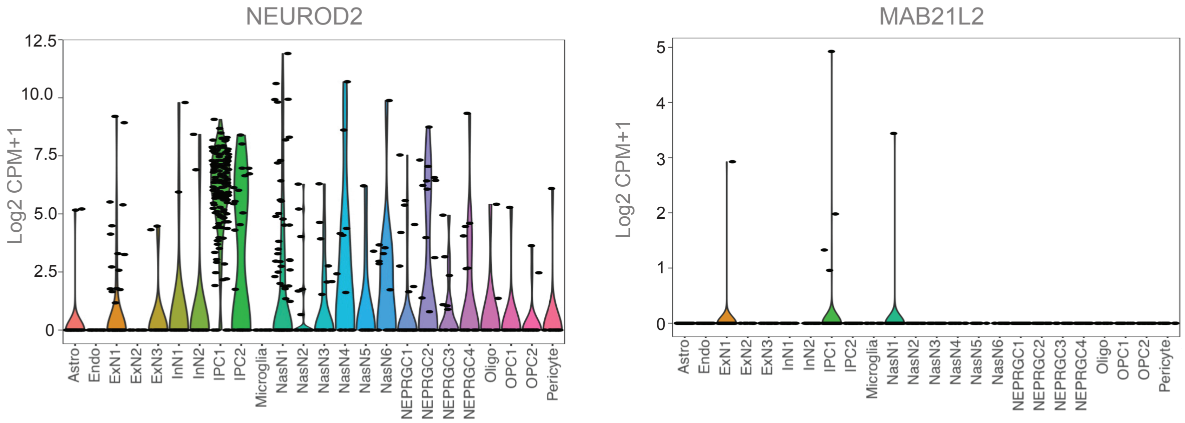


**Extended Data Figure 11.**

Violin plots showing NEUROD2 and MAB21L2 expression across cell types in the prenatal period, particularly in intermediate progenitor cells.

Astro, astrocyte; Endo, endothelial cell; ExN, excitatory neuron; InN, inhibitory neuron; IPC, intermediate progenitor cell; Microglia; NasN, nascent neuron; NEPRGC, neural epithelial progenitor/radial glial lineage; Oligo, oligodendrocyte; OPC, oligodendrocyte precursor cell; Pericyte. Analyzed transcriptomic dataset from PsychEncode.[51]
